# Interdependent Patient-Reported Outcome Patterns During Breast Cancer Pharmacotherapy: A Correlation-Based Analysis Using EORTC QLQ-C30 and QLQ-BR23

**DOI:** 10.64898/2026.02.10.26345961

**Authors:** Henry Sutanto, Merlyna Savitri, Een Hendarsih, Ami Ashariati

**Author notes:** **Correspondence:** Merlyna Savitri, MD and Henry Sutanto, MD, MSc, PhD, Department of Internal Medicine, Faculty of Medicine, Universitas Airlangga, Indonesia and.

## Abstract

**Background:** Quality-of-life (QoL) assessment is essential in breast cancer care, yet limited evidence describes how interrelated QoL domains change during pharmacotherapy. This study aimed to evaluate correlations among functional and symptom scales using the EORTC QLQ-C30 and QLQ-BR23, highlighting their ability to reveal multidimensional QoL patterns.

**Methods:** A prospective observational study was conducted in two second-referral hospitals in Indonesia, enrolling 106 female breast cancer patients. QoL was assessed before and after pharmacotherapy using QLQ-C30 and QLQ-BR23. Changes in scores (Δ) were computed, and interdomain relationships were analyzed using Spearman’s rho.

**Results:** Physical functioning correlated with role functioning (ρ = 0.55, *p* <0.001), emotial functioning (ρ = 0.33, *p* <0.001), and social functioning (ρ = 0.31, *p* = 0.002). Role and social functioning were likewise correlated (ρ = 0.32, *p* = 0.001), indicating that improvements across functional domains tended to occur in parallel. Symptom scales showed strong positive clustering, including fatigue with pain (ρ = 0.37, *p* <0.001), insomnia (ρ = 0.35, *p* <0.001), and systemic side effects (ρ = 0.48, *p* <0.001). Functional and symptom domains generally exhibited inverse relationships: physical functioning negatively correlated with fatigue (ρ = –0.40), pain (ρ = –0.43), both *p* <0.001, and systemic side effects (ρ = –0.26; p = 0.01).

**Conclusion:** The QLQ-C30 and QLQ-BR23 instruments effectively captured structured, clinically meaningful interdependencies. Functional improvements consistently aligned with symptom reductions, revealing coherent functional–symptom clustering. These findings underscore the sensitivity of QoL instruments to detect multidimensional patient-reported changes during breast cancer pharmacotherapy.

## Introduction

Breast cancer remains a major global health burden and is the most commonly diagnosed malignancy among women [1]. Advances in systemic therapy have led to substantial improvements in survival; however, these gains are often accompanied by complex physical, emotional, and social challenges that profoundly affect patients’ quality of life (QoL) [2,3]. As therapeutic strategies increasingly emphasize precision and patient-centered care, understanding how patients experience treatment—not only through clinical outcomes but also through the dynamic interplay of symptoms and functioning—has become a central priority in contemporary oncology. Patient-reported outcomes (PROs) now occupy a critical role in both clinical trials and routine care, providing insights that cannot be captured through clinician-reported toxicity or biomedical indicators alone [4]. Among the wide array of available PRO instruments, the European Organisation for Research and Treatment of Cancer (EORTC) QLQ-C30 and its breast cancer–specific module QLQ-BR23 stand as gold-standard tools for capturing multidimensional QoL in oncology [5,6]. These instruments are rigorously validated across cultures and languages, sensitive to change over time, and designed to profile distinct but interrelated domains of patient experience—including physical, emotional, cognitive, and social functioning, as well as treatment-related symptoms [7,8]. Their granular structure makes them uniquely suited not only for detecting changes within domains, but also for revealing relationships between domains, which may illuminate how patients adapt to treatment physiologically and psychosocially. Yet despite their widespread use, few studies leverage their full potential to explore how QoL domains interact with one another during the course of pharmacotherapy.

Emerging evidence suggests that QoL responses during cancer treatment do not occur in isolation; rather, functional and symptom domains often fluctuate in coordinated, patterned ways [9]. Traditional pre–post analyses may overlook these interdependencies, missing valuable information about the internal structure of patient experience. Correlation-based approaches offer an opportunity to map these interrelationships and identify clusters of domains that move together, potentially uncovering signatures of resilience, vulnerability, or treatment response [9]. Such insights may enhance clinical decision-making, enabling more targeted supportive care interventions and more refined prognostic assessments. However, data describing multidimensional correlation patterns among PRO domains in breast cancer—particularly within the context of pharmacotherapy in real-world low– and middle-income settings—remain scarce. Indonesia, where breast cancer tends to present at more advanced stages and where supportive care resources vary widely, represents an important setting to examine these relationships [10,11]. Understanding how functional and symptom domains shift together in this population may provide critical guidance for tailoring interventions and optimizing patient-centered care. In this study, we prospectively evaluated QoL before and after pharmacotherapy in breast cancer patients using the EORTC QLQ-C30 and QLQ-BR23 questionnaires. By analyzing changes across domains and their intercorrelations, we aimed to characterize the underlying structure of QoL dynamics during treatment. Our findings reveal coherent clustering of functional and symptom domains and demonstrate the value of EORTC instruments in capturing these complex patterns. This approach highlights how PROs can move beyond descriptive metrics to serve as integrative tools for understanding the lived experience of cancer therapy.

## Material and Methods

### Study design

This prospective observational study was conducted from January to October 2025 in two second-referral hospitals in Surabaya, Indonesia: Universitas Airlangga Hospital and Haji General Hospital. Both centers serve as major referral hubs for breast cancer management in East Java and provide comprehensive oncologic services, including surgery, systemic therapy, and supportive care. The study was designed to evaluate multidimensional changes in QoL during breast cancer pharmacotherapy through PROs using validated EORTC instruments.

### Ethical approval and informed consent

The study protocol received ethical clearance from the Ethics Committee of Universitas Airlangga Hospital (197/KEP/2024) and the Ethics Committee of Haji General Hospital (445/02/KOM.ETIK/2025). All procedures adhered to the Declaration of Helsinki. Written informed consent was obtained from all participants prior to enrollment.

### Participants and sampling method

Patients were recruited using consecutive sampling as they presented to oncology clinics in the participating hospitals. Adult women aged ≥18 years were eligible if they had a confirmed diagnosis of stage I–IV breast cancer established through concordant clinical findings, breast imaging (mammography or ultrasonography), and histopathology (fine-needle aspiration cytology or core biopsy). Additional inclusion criteria comprised availability or planned assessment of immunohistochemistry (IHC) markers (ER, PR, HER2, Ki-67), and current or planned pharmacotherapy in the form of chemotherapy, hormonal therapy, and/or targeted therapy. Patients were excluded if they had received ≥3 months of prior pharmacotherapy, had another active primary malignancy, were unable to communicate rationally, or had psychiatric conditions precluding reliable questionnaire completion.

### Data collection and clinical variables

Baseline demographic, clinical, and tumor characteristics were obtained from structured interviews and medical records. These included age, educational level, marital and employment status, comorbidities, disease stage, histopathology, tumor grade, IHC profile, and pharmacotherapy regimen. All data were collected by trained research staff to ensure consistency across study sites.

### Quality of life assessment

QoL was assessed using the Indonesian-validated versions of the EORTC QLQ-C30 and the breast cancer–specific module QLQ-BR23. Patients completed both questionnaires at two time points: (1) immediately before initiation of a treatment cycle, and (2) after completion of one full cycle of pharmacotherapy. These instruments evaluate multiple functional domains (physical, role, emotional, cognitive, and social functioning) and symptom domains (fatigue, pain, nausea/vomiting, insomnia, systemic side effects, breast and arm symptoms), enabling multidimensional assessment of treatment impact.

### Statistical analysis

Data distribution was assessed using the Shapiro–Wilk test; due to non-normality, QoL values were summarized as median with interquartile range (IQR). Change scores (Δ) were calculated to measure within-patient differences pre– and post-pharmacotherapy. Interdomain relationships among Δ-scores were analyzed using Spearman’s rho correlation coefficient. Correlation matrices were visualized using heatmaps and scatterplots to illustrate structural patterns among functional and symptom domains. Statistical analyses were conducted using SPSS version 26.0, with significance set at p < 0.05.

## Results

### Baseline characteristics of the study population

A total of 106 female breast cancer patients were enrolled, all of whom met the eligibility criteria and completed baseline assessments (**Table 1**). The mean age of participants was 51.92 ± 9.7 years, reflecting a predominantly middle-aged cohort. All participants were biologically female, consistent with the inclusion of clinically confirmed breast cancer cases. Educational attainment varied, with 1.9% having no formal education, 20.8% completing elementary school, 7.5% completing secondary school, 33.0% finishing high school, and 36.8% achieving higher education. Regarding employment status, 66.0% were unemployed or identified as housewives, while 34.0% were employed. Most participants were married (85.8%), whereas 7.5% were single and 6.6% were widowed. Comorbidities were present in 59.4% of patients, with hypertension being the most common (n = 32), followed by diabetes mellitus (n = 12), cardiovascular diseases (n = 16), respiratory diseases (n = 7), and other medical conditions (n = 37). The remaining 40.6% reported no comorbidities. Regarding prior breast cancer treatment, 69.8% had undergone at least one form of therapy before the assessment, including surgery (n = 60), pharmacotherapy (n = 49), or radiotherapy (n = 4). Meanwhile, 30.2% had not received any prior breast cancer–directed intervention at the time of baseline evaluation.

**Table 1.** Baseline characteristics of study population.

| Parameter | N = 106 |
| --- | --- |
| **Sex (n; %)** |  |
| Female | 106 (100.0%) |
| **Age in years (mean ± SD)** | 51.92 ± 9.7 |
| **Education (n; %)** |  |
| No formal education | 2 (1.9%) |
| Elementary | 22 (20.8%) |
| Secondary | 8 (7.5%) |
| High school | 35 (33.0%) |
| Higher education | 39 (36.8%) |
| **Employment Status (n; %)** |  |
| Unemployed / Housewife | 70 (66.0%) |
| Employed | 36 (34.0%) |
| **Marital Status (n; %)** |  |
| Single | 8 (7.5%) |
| Married | 91 (85.8%) |
| Widow | 7 (6.6%) |
| **Comorbidity (n; %)** |  |
| **None** | 43 (40.6%) |
| **Yes:** | 63 (59.4%) |
| Hypertension (n) | 32 |
| Diabetes mellitus (n) | 12 |
| Cardiovascular diseases (n) | 16 |
| Respiratory diseases (n) | 7 |
| Others (n) | 37 |
| **History of Breast Cancer Treatment (n; %)** |  |
| **None** | 32 (30.2%) |
| **Yes:** | 74 (69.8%) |
| Surgery (n) | 60 |
| Pharmacotherapy (n) | 49 |
| Radiotherapy (n) | 4 |

### Breast cancer profiles of the study population

The clinical and pathological characteristics of the 106 enrolled breast cancer patients are summarized in **Table 2**. The majority of patients presented with locally advanced disease (58.5%), followed by metastatic disease (28.3%) and early-stage breast cancer (13.2%). With respect to tumor histopathology, invasive ductal carcinoma (IDC) was the predominant subtype, identified in 84.0% of patients. Invasive lobular carcinoma (ILC) accounted for 4.7%, while 7.5% had mixed IDC/ILC histology. Rare histologic variants (e.g., mucinous carcinoma) were observed in 0.9% of cases, and 2.8% had insufficient data to determine subtype. Tumor grading revealed substantial heterogeneity: Grade II tumors were the most prevalent (35.8%), followed closely by Grade III tumors (40.6%), while Grade I tumors were less common (9.4%). Tumor grade could not be determined in 14.2% of samples due to incomplete pathology reporting. IHC profiling showed diverse molecular subtypes. Luminal B HER2-tumors were the most common subtype (26.4%), followed by Luminal B HER2+ (18.9%), Luminal A (9.4%), HER2-enriched (15.1%), HER2-low (6.6%), and triple-negative breast cancer (TNBC) (7.6%). IHC subtype was unavailable for 16.0% of participants. Regarding systemic treatment phase, 34.0% of patients were receiving neoadjuvant, 37.7% adjuvant, and 28.3% palliative pharmacotherapy. A wide range of chemotherapy, hormonal, and targeted regimens was documented, with docetaxel–carboplatin (n = 20), doxorubicin–cyclophosphamide combinations (n = 17), docetaxel–doxorubicin–cyclophosphamide (n = 12), and taxane/platinum–trastuzumab or its combinations among the most frequently administered. Multiple targeted and endocrine therapies, including trastuzumab, fulvestrant, aromatase inhibitors, and luteinizing hormone-releasing hormone (LHRH) analogs, were also used in smaller subsets of patients.

**Table 2.** Breast cancer profile of the study population.

| Parameter | N = 106 |
| --- | --- |
| **Disease Stage (n; %)** |  |
| Early | 14 (13.2%) |
| Locally advanced | 62 (58.5%) |
| Metastatic | 30 (28.3%) |
| **Histopathology (n; %)** |  |
| Invasive ductal carcinoma (IDC) | 89 (84.0%) |
| Invasive lobular carcinoma (ILC) | 5 (4.7%) |
| Mixed IDC/ILC | 8 (7.5%) |
| Others (e.g., mucinous) | 1 (0.9%) |
| Unknown | 3 (2.8%) |
| **Tumor Grade (n; %)** |  |
| Grade I | 10 (9.4%) |
| Grade II | 38 (35.8%) |
| Grade III | 43 (40.6%) |
| Unknown | 15 (14.2%) |
| **Immunohistochemistry (n; %)** |  |
| Luminal A | 10 (9.4%) |
| Luminal B HER2- | 28 (26.4%) |
| Luminal B HER2+ | 20 (18.9%) |
| HER2 *enriched* | 16 (15.1%) |
| HER2 *low* | 7 (6.6%) |
| TNBC | 8 (7.6%) |
| Unknown | 17 (16.0%) |
| **Pharmacotherapy Phase (n; %)** |  |
| Neoadjuvant | 36 (34.0%) |
| Adjuvant | 40 (37.7%) |
| Palliative | 30 (28.3%) |
| **Pharmacotherapy (n)** |  |
| Paclitaxel – Carboplatin | 8 |
| Docetaxel – Carboplatin | 20 |
| Docetaxel – Cyclophosphamide | 4 |
| Doxorubicin – Cyclophosphamide | 17 |
| Docetaxel – Doxorubicin – Cyclophosphamide | 12 |
| Docetaxel – Epirubicin – Cyclophosphamide | 5 |
| Epirubicin – Paclitaxel | 1 |
| Epirubicin – Docetaxel | 1 |
| Epirubicin – Carboplatin | 3 |
| Epirubicin – Cyclophosphamide – 5FU | 3 |
| Cyclophosphamide – Methotrexate – 5FU | 1 |
| Paclitaxel – Trastuzumab | 2 |
| Docetaxel – Trastuzumab | 2 |
| Paclitaxel – Carboplatin – Trastuzumab | 2 |
| Docetaxel – Carboplatin – Trastuzumab | 11 |
| Epirubicin – Docetaxel – Trastuzumab | 1 |
| Trastuzumab | 2 |
| Carboplatin – Gemcitabine | 1 |
| Paclitaxel – Gemcitabine | 1 |
| Docetaxel – Capecitabine | 1 |
| Vinorelbine | 3 |
| Eribulin | 1 |
| Leuprorelin | 1 |
| Tamoxifen | 2 |
| Fulvestrant | 1 |
| Letrozole | 3 |
| Goserelin | 7 |
| Bevacizumab | 2 |
| Asam zolendronik | 5 |

### Baseline QoL scores

Baseline QoL assessments using the EORTC QLQ-C30 and QLQ-BR23 demonstrated high functional status and variable symptom burden among the 106 participants (**Table 3**). The global health status/QoL score had a median of 83.3 [66.7–91.7], indicating generally favorable self-reported overall health prior to pharmacotherapy initiation. Across the QLQ-C30 functional scales, physical functioning showed a median of 93.3 [80.0–100.0], while role functioning, cognitive functioning, and social functioning all reached the scale maximum, with medians of 100.0 [100.0–100.0], 100.0 [83.3–100.0], and 100.0 [83.3–100.0], respectively. Emotional functioning demonstrated slightly lower scores, with a median of 83.3 [66.7–91.7]. Symptom scales revealed greater variability. Fatigue had a median score of 22.2 [0.0–44.4], while pain showed a median of 33.3 [16.7–50.0], representing one of the more prominent baseline symptoms. Insomnia exhibited a wide distribution, with a median of 33.3 [0.0–66.7]. Other symptoms such as nausea/vomiting, dyspnea, diarrhea, and constipation had median scores of 0.0, indicating minimal prevalence at baseline. Financial difficulty had a median of 0.0 [0.0–33.3], reflecting heterogeneous socioeconomic burden. On the QLQ-BR23 module, body image was high (median 100.0 [75.0–100.0]), whereas sexual functioning and sexual enjoyment scored lower (both 33.3 [0.0–33.3]). Future perspective showed moderate variability with a median of 66.7 [33.3–100.0]. Baseline breast-specific symptoms, including systemic side effects, breast symptoms, and arm symptoms, had median scores of 9.5 [4.8–19.1], 16.7 [6.3–27.1], and 11.1 [0.0–33.3], respectively, while upset by hair loss had a median of 0.0.

**Table 3.** Quality-of-life scale prior to pharmacotherapy.

| Parameter | Median [IQR]<sup>*</sup><br>(N = 106) |
| --- | --- |
| **Global health status** |  |
| Quality of life (QL2) | 83.3 [66.7 – 91.7] |
| **C30 – Functional Scale** |  |
| Physical function (PF2) | 93.3 [80.0 – 100.0] |
| Role function (RF2) | 100.0 [100.0 – 100.0] |
| Emotional function (EF) | 83.3 [66.7 – 91.7] |
| Cognitive function (CF) | 100.0 [83.3 – 100.0] |
| Social function (SF) | 100.0 [83.3 – 100.0] |
| **C30 – Symptom Scale** |  |
| Fatigue (FA) | 22.2 [0.0 – 44.4] |
| Nausea vomiting (NV) | 0.0 [0.0 – 0.0] |
| Pain (PA) | 33.3 [16.7 – 50.0] |
| Dyspnea (DY) | 0.0 [0.0 – 0.0] |
| Insomnia (SL) | 33.3 [0.0 – 66.7] |
| Loss of appetite (AP) | 0.0 [0.0 – 33.3] |
| Constipation (CO) | 0.0 [0.0 – 0.0] |
| Diarrhea (DI) | 0.0 [0.0 – 0.0] |
| Financial difficulty (FI) | 0.0 [0.0 – 33.3] |
| **BR23 – Functional Scale** |  |
| Body image (BRBI) | 100.0 [75.0 – 100.0] |
| Sexual function (BRSEF) | 33.3 [0.0 – 33.3] |
| Sexual enjoyment (BRSEE) | 33.3 [0.0 – 33.3] |
| Future perspective (BRFU) | 66.7 [33.3 – 100.0] |
| **BR23 – Symptom Scale** |  |
| Systemic side effect (BRST) | 9.5 [4.8 – 19.1] |
| Breast symptom (BRBS) | 16.7 [6.3 – 27.1] |
| Arm symptom (BRAS) | 11.1 [0.0 – 33.3] |
| Upset by hair loss (BRHL) | 0.0 [0.0 – 0.0] |

<sup>\*</sup> The data were not normally distributed based on the Shapiro–Wilk test ( $N < 100$ ), so the values are presented as median [IQR]

### Correlation patterns of Δ-QoL indicators

As shown in **Figure 1**, QL2 demonstrated strong associations with both functional status and symptom burden. Better overall QoL was significantly correlated with higher physical functioning (PF2; ρ = 0.331, *p* = 0.001), better role functioning (RF2; ρ = 0.407, *p* < 0.001), and a more positive future perspective (BRFU; ρ = 0.256, *p* = 0.010). Conversely, poorer QoL was significantly associated with higher fatigue (FA; ρ = –0.401, *p* <0.001), greater pain (PA; ρ = –0.404, *p* <0.001), increased appetite loss (AP; ρ = – 0.216, *p* = 0.032), and increased breast symptoms (BRST; ρ = –0.314, *p* = 0.002). No other functional, symptom, or breast cancer–specific subscales showed significant relationships with overall QoL. PF2 showed robust associations with multiple functional (**Figure 2**) and symptom domains (**Figure 3**). Higher PF2 scores were significantly associated with better role functioning (RF2; ρ = 0.549, *p* <0.001), emotional functioning (EF; ρ = 0.328, *p* = 0.001), and social functioning (SF; ρ = 0.306, *p* = 0.002). Improved PF2 was also linked to fewer symptoms, including fatigue (FA; ρ = –0.396, *p* <0.001), pain (PA; ρ = –0.434, *p* <0.001), dyspnea (DY; ρ = –0.310, *p* = 0.002), sleep disturbance (SL; ρ = –0.335, *p* = 0.001), appetite problems (AP; ρ = –0.400, *p* <0.001), and constipation (CO; ρ = –0.250, *p* = 0.012). Among breast cancer–specific domains, higher PF2 correlated positively with systemic therapy side-effect functioning (BRSEF; ρ = 0.208, *p* = 0.039) and sexual enjoyment (BRSEE; ρ = 0.230, *p* = 0.022), while being inversely associated with breast-specific symptoms (BRST; ρ = –0.259, *p* = 0.010). Finally, PF2 was positively associated with overall QoL (QL2; ρ = 0.331, *p* = 0.001). These findings indicate that better physical functioning aligns consistently with both enhanced psychosocial functioning and reduced symptom burden. Meanwhile, RF2 exhibited a strong pattern of associations with multiple functional and symptom domains. Higher RF2 scores were significantly correlated with better physical functioning (PF2; ρ = 0.549, *p* <0.001), improved social functioning (SF; ρ = 0.316, *p* = 0.001), and higher overall QoL (QL2; ρ = 0.407, *p* <0.001). Conversely, better role functioning was associated with lower symptom burden, reflected by significant inverse correlations with fatigue (FA; ρ = –0.498, *p* <0.001), pain (PA; ρ = –0.372, *p* <0.001), sleep disturbance (SL; ρ = –0.332, *p* = 0.001), appetite problems (AP; ρ = –0.412, *p* <0.001), and diarrhea (DI; ρ = –0.248, *p* = 0.013). Among breast cancer–specific domains, RF2 was negatively correlated with breast symptoms (BRST; ρ = –0.413, *p* <0.001) and arm symptoms (BRAS; ρ = –0.224, *p* = 0.026). These findings highlight that better role functioning is consistently aligned with enhanced global QoL and reduced multidimensional symptom burden. EF demonstrated a selective pattern of significant associations with both functional and symptom domains. Higher EF scores were significantly correlated with better physical functioning (PF2; ρ = 0.328, *p* = 0.001) and improved future perspective (BRFU; ρ = 0.273, *p* = 0.006), indicating that stronger emotional functioning aligned with enhanced overall outlook and physical capability. Conversely, emotional functioning was inversely associated with several symptom indicators, including pain (PA; ρ = –0.207, *p* = 0.040) and sleep disturbance (SL; ρ = – 0.225, *p* = 0.025), suggesting that higher emotional well-being corresponded with lower pain perception and fewer sleep-related problems. No other domains demonstrated significant relationships with EF. Interestingly, CF did not show any statistically significant correlations with other functional domains, symptom scales, or breast cancer–specific subscales, while SF demonstrated significant associations with several functional and symptom domains. Higher social functioning was positively correlated with better physical functioning (PF2; ρ = 0.306, *p* = 0.002) and improved role functioning (RF2; ρ = 0.316, *p* = 0.001), indicating that enhanced social engagement aligned with better overall functional status. Conversely, SF was negatively associated with fatigue (FA; ρ = –0.281, *p* = 0.005) and pain (PA; ρ = –0.207, *p* = 0.040), suggesting that higher social functioning corresponded with lower symptom burden. No other significant correlations were observed.

**Figure 1.**
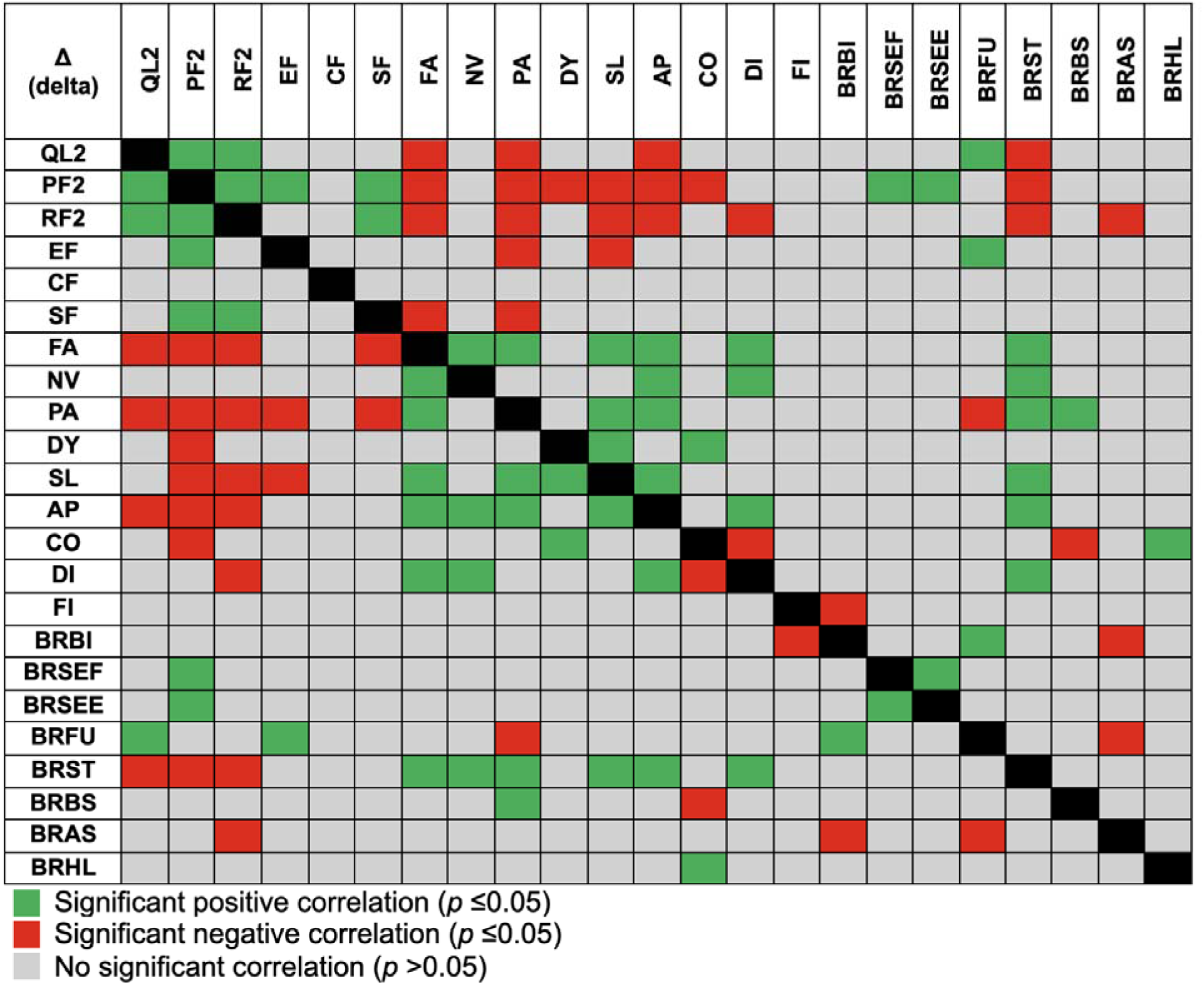
Spearman’s rho correlation heatmap of the changes (Δ) in QoL parameters among breast cancer patients before and after pharmacotherapy.

**Figure 2.**
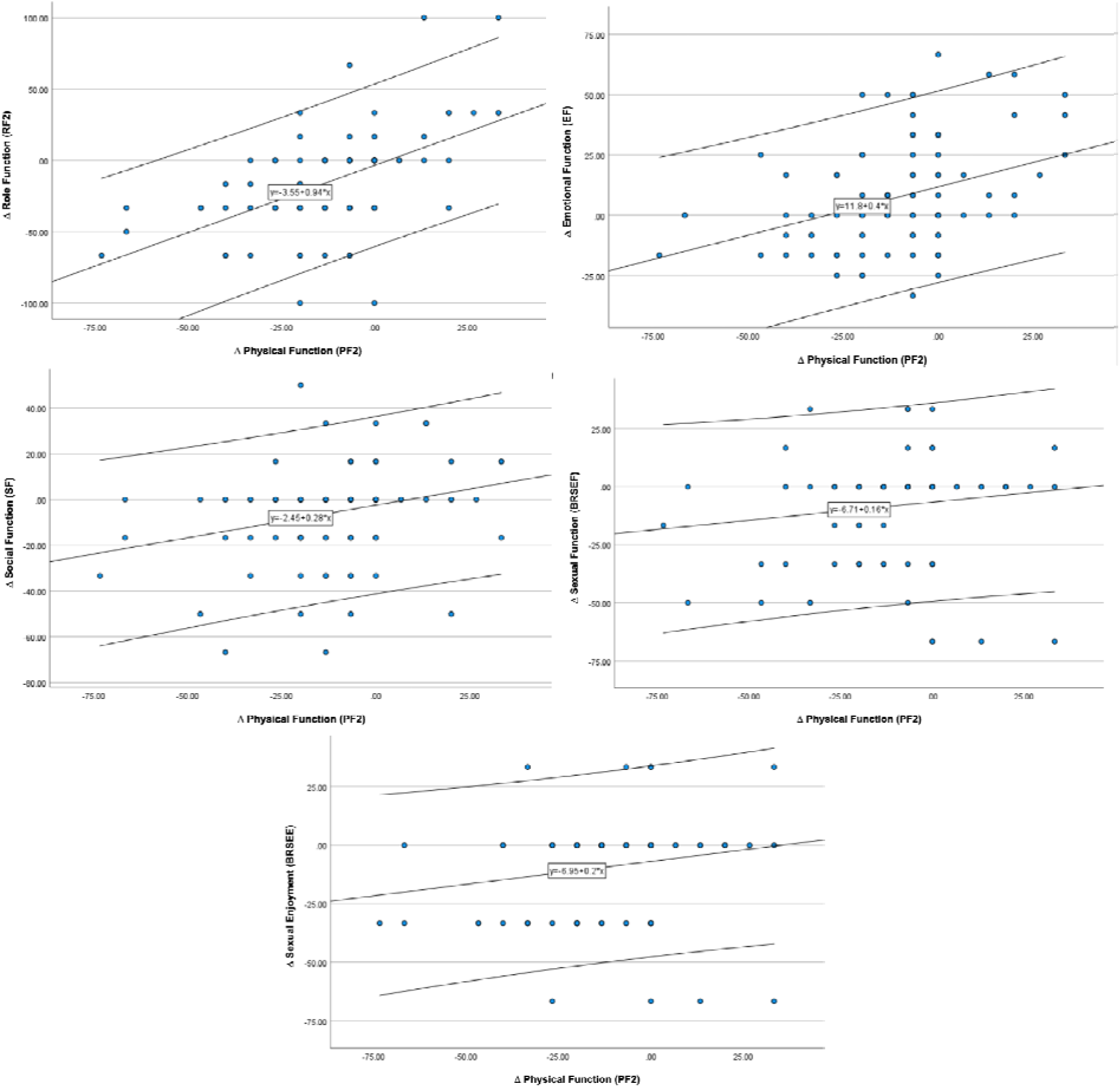
Scatter plot illustrating the significant positive correlation between PF2 and other QoL parameters.

**Figure 3.**
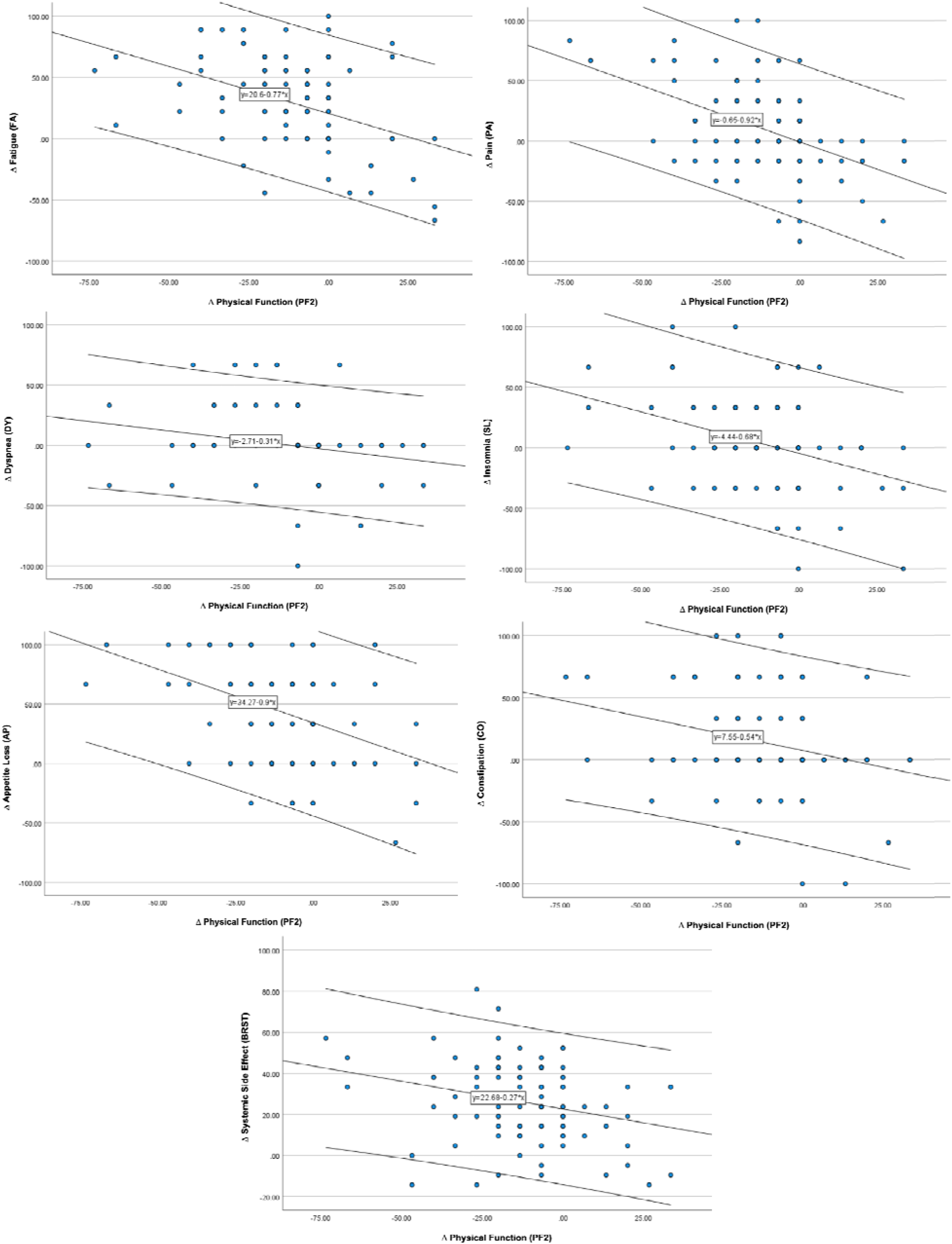
Scatter plot illustrating the significant negative correlation between PF2 and other QoL parameters.

In the symptom domain, FA displayed a wide range of significant associations across both functional and symptom domains. Higher fatigue levels were strongly associated with lower physical functioning (PF2; ρ = –0.396, *p* <0.001), poorer role functioning (RF2; ρ = –0.498, *p* <0.001), and reduced social functioning (SF; ρ = – 0.281, *p* = 0.005). Fatigue was also negatively correlated with overall QoL (QL2; ρ = – 0.401, *p* <0.001), highlighting its broad detrimental impact. Conversely, FA was positively correlated with multiple symptom burdens, including nausea/vomiting (NV; ρ = 0.331, *p* = 0.001), pain (PA; ρ = 0.367, *p* <0.001), sleep disturbance (SL; ρ = 0.350, *p* <0.001), appetite loss (AP; ρ = 0.527, *p* <0.001), and diarrhea (DI; ρ = 0.240, *p* = 0.017). Among breast cancer–specific domains, FA was significantly associated with greater breast symptom burden (BRST; ρ = 0.482, *p* <0.001). Collectively, these findings demonstrate that fatigue is centrally linked to both impaired functioning and heightened multisystem symptom severity. NV demonstrated a focused pattern of significant associations with several key symptom domains. Higher nausea and vomiting scores correlated positively with greater fatigue (FA; ρ = 0.331, *p* = 0.001), appetite loss (AP; ρ = 0.446, *p* <0.001), and diarrhea (DI; ρ = 0.257, *p* = 0.010), indicating clustering of gastrointestinal and systemic symptom burdens. Additionally, NV was positively associated with breast symptoms (BRST; ρ = 0.314, *p* = 0.002), suggesting a link between nausea/vomiting and heightened disease– or treatment-related breast discomfort. No significant correlations were observed with functional domains or other breast cancer–specific scales. PA showed a broad range of significant associations with both functional outcomes and symptom burden. Higher pain levels were significantly associated with lower physical functioning (PF2; ρ = –0.434, *p* <0.001), poorer role functioning (RF2; ρ = –0.372, *p* <0.001), and reduced emotional and social functioning (EF and SF; both ρ = –0.207, *p* = 0.040). Pain also correlated negatively with overall QoL (QL2; ρ = –0.404, *p* <0.001) and with future perspective (BRFU; ρ = –0.213, *p* = 0.034). Conversely, PA demonstrated strong positive correlations with multiple symptom domains, including fatigue (FA; ρ = 0.367, *p* <0.001), sleep disturbance (SL; ρ = 0.351, *p* <0.001), appetite loss (AP; ρ = 0.229, *p* = 0.022), and diarrhea (DI; ρ = 0.240, *p* = 0.017). Within breast cancer–specific scales, higher pain was significantly associated with greater breast symptoms (BRST; ρ = 0.329, *p* = 0.001) and breast-related side effects (BRBS; ρ = 0.238, *p* = 0.018). Overall, pain emerged as a central symptom strongly linked to both impaired functioning and increased multisystem symptomatology. DY showed a focused pattern of significant associations. Higher dyspnea scores were significantly associated with lower physical functioning (PF2; ρ = –0.310, *p* = 0.002), indicating that worsening breathlessness corresponded closely with reduced mobility and functional capacity. Dyspnea was also positively correlated with sleep disturbance (SL; ρ = 0.226, *p* = 0.024) and constipation (CO; ρ = 0.218, *p* = 0.030), suggesting clustering with other distressing symptoms. No significant associations were observed with role, emotional, cognitive, or social functioning, nor with breast cancer–specific domains. SL showed significant associations across multiple functional and symptom domains. Higher sleep disturbance was strongly associated with lower physical functioning (PF2; ρ = –0.335, *p* = 0.001), poorer role functioning (RF2; ρ = –0.332, *p* = 0.001), and reduced emotional functioning (EF; ρ = –0.225, *p* = 0.025). Conversely, sleep disturbance correlated positively with several symptom indicators, including fatigue (FA; ρ = 0.350, *p* <0.001), pain (PA; ρ = 0.351, *p* <0.001), dyspnea (DY; ρ = 0.226, *p* = 0.024), and appetite loss (AP; ρ = 0.276, *p* = 0.006). Within breast cancer–specific domains, SL was positively associated with breast symptoms (BRST; ρ = 0.285, *p* = 0.004). No other significant relationships were identified.

AP demonstrated strong associations across functional, symptom, and breast cancer–specific domains. Greater appetite loss was significantly correlated with poorer physical functioning (PF2; ρ = –0.400, *p* <0.001) and role functioning (RF2; ρ = –0.412, *p* <0.001), as well as lower overall QoL (QL2; ρ = –0.216, *p* = 0.032). Appetite loss was also strongly linked to higher symptom burden, including fatigue (FA; ρ = 0.527, *p* <0.001), nausea/vomiting (NV; ρ = 0.446, *p* <0.001), pain (PA; ρ = 0.229, *p* = 0.022), sleep disturbance (SL; ρ = 0.276, *p* = 0.006), and diarrhea (DI; ρ = 0.336, *p* = 0.001). Among breast cancer–specific scales, AP was strongly associated with breast symptoms (BRST; ρ = 0.520, *p* <0.001). No other significant correlations were observed. CO demonstrated several significant associations spanning functional, symptom, and breast cancer–specific domains. Higher constipation scores were associated with lower physical functioning (PF2; ρ = –0.250, *p* = 0.012) and with reduced gastrointestinal well-being, as reflected by a negative correlation with diarrhea (DI; ρ = –0.285, *p* = 0.004). Conversely, constipation correlated positively with dyspnea (DY; ρ = 0.218, *p* = 0.030), suggesting clustering with other discomfort-related symptoms. Among breast cancer–specific scales, CO showed significant associations with greater breast-related side effects (BRBS; ρ = –0.203, *p* = 0.044) and increased breast/arm discomfort (BRHL; ρ = 0.230, *p* = 0.022). No other significant relationships were observed. DI demonstrated several significant associations with both functional and symptom domains. Higher diarrhea scores were associated with poorer role functioning (RF2; ρ = –0.248, *p* = 0.013) and with lower gastrointestinal well-being, as reflected by a negative correlation with constipation (CO; ρ = –0.285, *p* = 0.004). Conversely, diarrhea was positively associated with multiple symptom indicators, including fatigue (FA; ρ = 0.240, *p* = 0.017), nausea/vomiting (NV; ρ = 0.257, *p* = 0.010), and appetite loss (AP; ρ = 0.336, *p* = 0.001). Within breast cancer–specific scales, DI correlated significantly with greater breast symptoms (BRST; ρ = 0.333, *p* = 0.001). No other meaningful associations were observed. FI only showed a single significant association within the breast cancer–specific domains. Lower financial impact scores (indicating less financial difficulty) were associated with better body image (BRBI; ρ = – 0.256, *p* = 0.011). No significant correlations were observed with global functioning, symptom burden, or other breast cancer–specific scales.

In the breast-cancer-specific domain, BRBI demonstrated a focused set of significant associations with both financial and breast cancer–specific domains. Lower body image scores were associated with greater financial difficulty (FI; ρ = –0.256, *p* = 0.011) and higher arm symptom burden (BRAS; ρ = –0.230, *p* = 0.022). Conversely, better body image was positively associated with a more favorable future perspective (BRFU; ρ = 0.207, *p* = 0.040). No significant correlations were observed with functional scales, core symptom domains, or other breast cancer–specific subscales. BRSEF showed two significant associations. Higher systemic therapy side-effect scores were modestly associated with lower physical functioning (PF2; ρ = 0.208, *p* = 0.039). The strongest relationship was observed with sexual enjoyment (BRSEE; ρ = 0.874, *p* <0.001), reflecting a very strong internal correlation between breast cancer–specific symptom subscales. No other functional, symptom, or breast cancer–specific domains demonstrated significant associations with BRSEF. BRSEE also showed two significant associations. Higher sexual enjoyment was modestly associated with better physical functioning (PF2; ρ = 0.230, *p* = 0.022). The strongest correlation was with systemic therapy side effects (BRSEF; ρ = 0.874, *p* < 0.001), reflecting substantial overlap between these breast cancer–specific subscales. No significant relationships were observed with functional scores, symptom domains, or other breast cancer–specific measures. Whilst, BRFU showed several significant associations across functional and breast cancer–specific domains. Higher future perspective was strongly associated with better emotional functioning (EF; ρ = 0.273, *p* = 0.006) and more positive body image (BRBI; ρ = 0.207, *p* = 0.040). BRFU was also positively associated with overall QoL (QL2; ρ = 0.256, *p* = 0.010). Conversely, a more negative future perspective was linked to greater pain levels (PA; ρ = –0.213, *p* = 0.034) and higher arm symptom burden (BRAS; ρ = –0.250, *p* = 0.012). No other significant correlations were observed.

BRST showed extensive associations with both functional and symptom domains. Higher breast symptom burden was significantly associated with lower physical functioning (PF2; ρ = –0.259, *p* = 0.010), poorer role functioning (RF2; ρ = – 0.413, *p* <0.001), and reduced overall QoL (QL2; ρ = –0.314, *p* = 0.002). Conversely, BRST was strongly positively correlated with multiple symptoms, including fatigue (FA; ρ = 0.482, *p* < 0.001), nausea/vomiting (NV; ρ = 0.314, *p* = 0.002), pain (PA; ρ = 0.329, *p* = 0.001), sleep disturbance (SL; ρ = 0.285, *p* = 0.004), appetite loss (AP; ρ = 0.520, *p* <0.001), and diarrhea (DI; ρ = 0.333, *p* = 0.001). No other functional or breast cancer–specific domains demonstrated significant associations. BRBS demonstrated only two significant associations. Higher breast-specific symptom scores were modestly correlated with increased pain (PA; ρ = 0.238, *p* = 0.018), indicating that greater breast symptoms coincide with higher pain burden. Conversely, breast symptoms were negatively associated with constipation (CO; ρ = –0.203, *p* = 0.044). BRAS demonstrated several significant negative associations within functional and breast cancer–specific domains. Higher arm symptom burden was associated with poorer role functioning (RF2; ρ = –0.224, *p* = 0.026), reduced body image (BRBI; ρ = –0.230, *p* = 0.022), and a more negative future perspective (BRFU; ρ = –0.250, *p* = 0.012). Finally, BRHL showed only a single significant association. Higher breast and hormonal symptom burden was modestly correlated with greater constipation (CO; ρ = 0.230, *p* = 0.022). No significant associations were observed with functional scores, general symptom domains, or other breast cancer–specific subscales.

## Discussion

Overall, this prospective study provides a comprehensive evaluation of QoL dynamics among Indonesian breast cancer patients undergoing pharmacotherapy and demonstrates how multidimensional PROs can reveal structured patterns of functional and symptom changes within a single treatment cycle. Baseline assessments showed that most patients entered treatment with relatively high functional status yet notable variability in symptom intensity. Using the EORTC QLQ-C30 and QLQ-BR23 instruments, we analyzed the change scores across all domains and identified a distinct correlation architecture: functional domains displayed consistent positive associations with each other, symptom domains formed their own cohesive cluster of positive correlations, and the majority of cross-domain relationships were significantly negative. These patterns were visualized clearly in the correlation heatmap, which showed aligned trajectories within functional scales and within symptom scales, while **Figures 2 and 3** further illustrated that changes in physical functioning were positively associated with other functional improvements and negatively associated with several treatment-related symptoms.

### Baseline patient characteristics highlight diverse socioclinical backgrounds

The baseline characteristics of the study population reveal important contextual factors that shape how QoL patterns should be interpreted in this cohort. The mean age of participants—approximately 52 years—aligns with the epidemiologic profile of breast cancer in many low– and middle-income countries (LMICs), where patients tend to present at a younger age compared with Western populations [12]. This relatively younger demographic often carries distinct psychosocial and economic burdens, particularly among women balancing employment, caregiving responsibilities, and household management [13]. The high proportion of unemployed or housewife participants (66%) further highlights these socioeconomic vulnerabilities and may influence domains such as role functioning, social functioning, and financial difficulty. These structural, non-biological determinants of wellbeing are known to modulate patient-reported outcomes and may partly explain the heterogeneity observed in several baseline QoL scores. Educational attainment showed broad variability, with substantial representation from high school and higher education levels. Educational status has been repeatedly associated with health literacy, treatment adherence, and the ability to navigate complex cancer care pathways [14,15]. In this cohort, the relatively balanced distribution across education levels suggests a diverse range of patient capacities for understanding treatment trajectories, which may influence emotional functioning, subjective symptom reporting, and future perspective. Similarly, the high proportion of married participants (85.8%) indicates that most patients entered treatment with potential social support structures in place. Previous studies have shown that marital support can significantly impact QoL trajectories, affecting domains such as emotional functioning, fatigue perception, and overall resilience during therapy [16–18].

Comorbid conditions were present in over half of the patients, with hypertension and cardiovascular diseases being the most common. These comorbidities introduce additional physiological stressors that may influence the symptom profiles of patients undergoing chemotherapy or endocrine therapy. They also present a background of chronic medication use that could intersect with cancer treatment side effects, particularly fatigue, dyspnea, or insomnia. The presence of respiratory diseases and other chronic conditions in smaller subsets of patients underscores the complex clinical landscape in which pharmacotherapy is administered and may partly shape the correlation patterns observed between functional decline and symptom escalation. Notably, nearly 70% of the cohort had already undergone at least one cancer-directed intervention—most commonly surgery—while a substantial proportion had also received previous pharmacotherapy. These pre-treatment exposures introduce baseline variability in body image, fatigue levels, arm symptoms, and emotional readiness, all of which were captured in the initial QoL assessment. Patients who had undergone surgery may exhibit improved tumor-related symptoms but also experience lingering postoperative discomfort or functional limitations, particularly in arm mobility [19]. In contrast, patients without prior treatment may report higher levels of anxiety or uncertainty, affecting emotional functioning or future perspective. The diversity in treatment history strengthens the external validity of the study but also underscores the importance of analyzing QoL changes relative to individualized clinical trajectories.

### Clinical and molecular diversity of breast cancer presentation

The clinical and pathological profile of this cohort reflects the prevailing patterns of breast cancer presentation in Indonesia and many other LMICs, where delayed diagnosis and heterogeneous access to oncology services contribute to more advanced disease at presentation [11]. Over half of the patients in this study presented with locally advanced disease (58.5%), and nearly one-third had metastatic disease at baseline. This degree of disease burden has direct implications for the interpretation of QoL trajectories, as advanced stage is strongly associated with higher symptom load, greater functional limitations, and increased psychological distress [20]. Moreover, patients undergoing pharmacotherapy at later stages often have more complex care needs, and their QoL responses may be more sensitive to fluctuations in treatment intensity and symptom burden compared with early-stage patients [21,22]. Histopathologically, the predominance of IDC at 84% is consistent with global and regional epidemiologic data, underscoring IDC as the most common breast cancer subtype across diverse populations. The relatively small proportion of invasive lobular carcinoma, mixed histologies, and rare variants reflects the typical distribution seen in real-world oncology practice. While histologic subtype itself may influence symptom perception or functional impairment, its more important role in this analysis lies in its intersection with treatment choices and molecular profiling. The notable proportion of tumors graded as II (35.8%) and III (40.6%) indicates an overall moderate-to-high proliferation phenotype in this cohort. Higher-grade tumors are often associated with more aggressive biological behavior and, consequently, more intensive chemotherapy regimens, which can amplify treatment-related symptoms captured in the QLQ-C30 and QLQ-BR23 symptom scales. The IHC profile further highlights the biological diversity of the cohort. Luminal B HER2– and Luminal B HER2+ tumors together accounted for a large proportion of cases, aligning with the increasingly recognized predominance of Luminal B subtypes in Asian populations [23,24]. These subtypes often necessitate multi-agent chemotherapy, endocrine therapy, or a combination thereof, all of which influence the functional and symptom domains measured. The presence of HER2-enriched tumors (15.1%) and HER2-low tumors (6.6%) reflects evolving diagnostic practices, including more widespread availability of HER2 testing. HER2-low categorization, in particular, is clinically relevant given emerging links to antibody–drug conjugate (ADC) sensitivity. Meanwhile, the triple-negative subgroup accounted for 7.6% of cases—smaller than some regional reports but still clinically significant given the aggressive nature and limited targeted therapies available for TNBC. Patients within this subgroup may experience more intensive cytotoxic therapy and, therefore, higher symptom variability. The 16% of cases with unknown IHC status likely reflect real-world gaps in diagnostic availability or incomplete reporting, a common challenge in resource-variable settings that may complicate QoL interpretation.

The distribution of treatment phases—neoadjuvant (34%), adjuvant (37.7%), and palliative (28.3%)—underscores the broad therapeutic spectrum represented in this cohort. Patients receiving neoadjuvant therapy may present with substantial baseline tumor-related symptoms, while those receiving adjuvant therapy may experience cumulative toxicity following surgery. Palliative patients, in contrast, often have disease-driven symptom burdens that interact dynamically with treatment side effects. These distinctions are crucial when interpreting Δ-QoL correlations, as the magnitude and direction of change may differ depending on baseline disease status, symptom trajectory, and treatment intent. The breadth of pharmacotherapy regimens documented—from anthracycline-based combinations to taxane–carboplatin regimens, HER2-targeted therapies, endocrine therapy, and targeted agents—highlights the heterogeneity of systemic treatment exposure [25,26]. Such diversity strengthens the ecological validity of the study but also introduces multiple potential sources of variation in symptom and function changes. Anthracyclines and taxanes, for instance, have well-characterized profiles of fatigue, neuropathy, nausea, and cytopenias, while endocrine therapies may influence cognitive, emotional, or sexual functioning [27–29]. This therapeutic heterogeneity mirrors contemporary clinical practice and enriches the QoL signal captured by the EORTC instruments, as different regimens may selectively influence distinct QoL domains.

### Baseline QoL scores reveal strong functioning and heterogeneous symptoms

The baseline QoL scores presented in **Table 3** offer critical insight into the functional and symptomatic landscape with which patients entered pharmacotherapy, and they provide an essential reference point for understanding subsequent changes. Overall, the cohort demonstrated unexpectedly high baseline functional scores across multiple domains. Physical functioning, cognitive functioning, role functioning, and social functioning exhibited medians at or near the upper limits of the scale. These findings may reflect the demographic composition of the cohort—patients with a mean age in the early fifties who may retain substantial physical reserve—and the possibility that many were evaluated at a point where functional decline had not yet manifested despite the presence of cancer. High social functioning scores may also reflect strong family and community support structures common in Indonesian cultural settings, as well as high marriage rates within the cohort [30]. Emotional functioning, while still high overall, showed more variability than other functional domains, with a broader interquartile range. This pattern is consistent with the psychological complexity associated with receiving a cancer diagnosis and entering systemic therapy. Emotional scores are often influenced by anxiety, uncertainty, and treatment anticipation rather than physical illness alone. The presence of such variability is clinically meaningful, as emotional functioning is one of the domains most likely to show dynamic changes during treatment and may be particularly sensitive to symptom fluctuations, as later correlation analyses suggest [31].

Symptom burden at baseline was heterogeneous, with several symptoms reported at low frequencies yet others showing notable variability. Fatigue and pain were the most prominent baseline symptoms, reflected by moderate median values and broad distribution ranges. These two symptoms are commonly reported even before treatment initiation and may stem from tumor-related metabolic and inflammatory processes, pre-existing comorbidities, or psychological stress [32,33]. Insomnia, which also showed a wide interquartile range, may similarly reflect both physiological and emotional contributors. In contrast, nausea/vomiting, dyspnea, diarrhea, and constipation appeared minimal at baseline, reinforcing that most patients had not yet experienced treatment-related toxicity. This absence of pharmacotherapy-induced symptoms establishes a clean baseline against which treatment effects can be detected.

Breast cancer–specific QoL scales exhibited additional distinctions. Body image scores were high in most patients, which could be attributed to the fact that 30% had not yet undergone surgery and many others had already adapted to postoperative changes. However, domains related to sexual functioning and sexual enjoyment showed markedly low baseline scores. These findings are consistent with global trends demonstrating that sexuality is often affected early in the cancer trajectory, influenced by psychological distress, relationship changes, cultural expectations, and hormonal alterations associated with the disease itself [34,35]. Future perspective also showed considerable variability, reflecting diverse levels of optimism, uncertainty, or coping readiness among patients. Symptoms specific to the breast cancer module—including breast symptoms, arm symptoms, and systemic side effects—were present but generally low, suggesting that local disease-related discomfort existed in only a subset of patients. Upset by hair loss scored at zero for many patients, which is expected before chemotherapy initiation and provides a meaningful contrast for post-treatment assessments. Taken together, these baseline QoL findings delineate a cohort beginning pharmacotherapy with high functional capacity, moderate emotional variability, and selective symptom burdens that are likely driven by tumor biology rather than treatment toxicity. This configuration establishes a robust foundation for interpreting Δ-QoL correlations, as changes observed after pharmacotherapy can be more confidently attributed to treatment-related effects rather than pre-existing limitations. Moreover, the heterogeneity observed across several symptom scales underscores the importance of using multidimensional patient-reported outcome tools such as the EORTC QLQ-C30 and QLQ-BR23 to capture diverse patient experiences at baseline and track nuanced changes over time.

### Correlation matrix reveals structured QoL interdependencies

The correlation matrix presented in **Figure 1** provides one of the most informative insights into the QoL dynamics observed in this study, revealing that patient-reported outcomes during pharmacotherapy do not change in isolation but instead evolve in tightly interconnected patterns. The emergence of two distinct clusters—one comprising functional domains and the other comprising symptom domains—supports the conceptual view that functional capacity and symptom burden represent two major, interacting dimensions of the patient experience during cancer treatment [36]. The strong positive correlations observed within each cluster suggest that changes within a given domain are not domain-specific phenomena but part of broader, domain-wide shifts. For example, when one aspect of daily functioning improves or deteriorates, other functional abilities tend to change in the same direction. This interdependence reinforces earlier psychometric evaluations of the QLQ-C30, which have consistently shown that functional items load cohesively on shared latent constructs, but our findings extend this understanding by demonstrating how these constructs behave dynamically during active treatment. Equally notable is the pattern of correlations within the symptom cluster, which showed consistent co-movement among prominent symptoms such as fatigue, pain, insomnia, and breast– or arm-related discomfort. These symptoms are often driven by shared biological mechanisms—such as inflammation, neuroendocrine stress responses, or cumulative toxicity from pharmacotherapy—and the strong positive correlations observed here are consistent with prior studies demonstrating symptom clustering in oncology populations. Importantly, the close alignment of treatment-related symptoms with breast-cancer–specific symptoms from the BR23 module highlights the value of using both generic and disease-specific scales concurrently. Together, these scales allowed the analysis to detect symptom constellations that might otherwise remain obscured if only one instrument were used.

Perhaps the most clinically relevant finding in **Figure 1** is the broad band of significant negative correlations between the functional and symptom domains. This inverse relationship reflects the intuitive clinical phenomenon that as symptoms intensify—whether due to disease burden or treatment toxicity—functional abilities tend to decline. However, the strength and consistency of these correlations across nearly all functional–symptom pairings suggest a deeper structural relationship: functional decline and symptom escalation may reflect two sides of the same underlying process. This pattern supports contemporary models of QoL in oncology that emphasize the reciprocal feedback between physical capacity and symptom distress [37]. The fact that these negative correlations appear broadly and not selectively implies that physical, emotional, social, and cognitive functioning may be comparably vulnerable to symptom burden, underscoring the holistic impact of pharmacotherapy on patients’ lived experience. The breast cancer–specific functional domains, particularly body image and future perspective, demonstrated an intermediate pattern of associations. Rather than clustering exclusively with either functional or symptom domains, they showed meaningful correlations with both. This cross-domain positioning suggests that breast cancer–specific concerns are shaped by an interplay between functional capacity and symptom burden, making them sensitive and integrative indicators of treatment impact [38]. By contrast, domains such as sexual functioning, sexual enjoyment, and upset by hair loss demonstrated minimal connectivity to other scales, consistent with prior literature showing that their trajectories often follow distinct psychosocial pathways, independent of short-term pharmacologic toxicity. Overall, the structure of correlations in **Figure 1** highlights the capacity of the EORTC QLQ-C30 and QLQ-BR23 to detect not only changes in individual domains but also the relational architecture of these changes. This multidimensional sensitivity is a core strength of PRO measures and is particularly valuable in heterogeneous real-world populations such as ours. The coherent, clinically interpretable clustering patterns further support the use of these validated instruments for monitoring treatment response, guiding supportive care, and understanding the lived complexities of breast cancer therapy in resource-variable settings.

### Improvement in physical functioning coincides with gains across multiple QoL domains

**Figure 2** provides a clear visualization of the positive correlations between changes in physical functioning (ΔPF2) and several other QoL domains, underscoring the interconnected nature of patient-reported functioning during breast cancer pharmacotherapy. The consistent upward trajectories observed across these scatter plots suggest that when patients experience an improvement in their physical functioning following treatment, other dimensions of functioning frequently improve alongside it. This pattern aligns with established models of multidimensional functioning in oncology, in which physical capacity is closely tied to emotional resilience, role functioning, and the ability to maintain social roles [39,40]. What Figure 2 adds, however, is evidence that these relationships are not static but dynamically responsive to even a single cycle of pharmacotherapy, highlighting the sensitivity of patient-reported outcomes to early and subtle shifts in wellbeing. The positive association between ΔPF2 and role functioning reflects the logical linkage between physical capability and the ability to carry out daily tasks or fulfill occupational and familial responsibilities. Similarly, the upward trends involving emotional functioning suggest that improvements in physical functioning may coincide with reductions in treatment-related distress. These associations resonate with patient narratives in clinical practice, where individuals often report better mood, clarity, and engagement when their bodies feel more capable [41]. From a psychometric perspective, the clustering of these improvements reinforces the discriminant yet interconnected structure of the QLQ-C30 functional scales, supporting the instrument’s ability to capture distinct but mutually influential aspects of functioning. The positive correlations between ΔPF2 and breast cancer–specific functional domains such as sexual function and sexual enjoyment add another important dimension to this interpretation. These relationships demonstrate how improvements in physical functioning can ripple across broader aspects of psychosocial wellbeing, reinforcing the idea that QoL improvements in oncology often occur as part of a coherent pattern rather than isolated domain-specific events. Overall, Figure 2 demonstrates that physical functioning operates as an integral node within the broader network of QoL domains, not by virtue of centrality in a statistical sense, but by illustrating that changes in physical functioning frequently occur in parallel with improvements across multiple functional and psychosocial dimensions. These findings highlight the utility of the EORTC QLQ-C30 and QLQ-BR23 in capturing early, clinically relevant shifts in patient wellbeing and underscore the importance of monitoring functional recovery as part of comprehensive supportive care during pharmacotherapy.

### Worsening symptoms correspond with reductions in physical functioning

**Figure 3** illustrates the significant negative correlations between changes in ΔPF2 and multiple symptom domains, offering critical insight into how symptom burden and functional status interact during pharmacotherapy. The consistent downward slopes across the scatter plots indicate that worsening symptoms—particularly fatigue, pain, insomnia, and systemic side effects—are closely aligned with reductions in physical functioning. This inverse pattern is well-recognized clinically but is rarely captured in such a structured, data-driven manner. The EORTC QLQ-C30 and QLQ-BR23 instruments make it possible to visualize and quantify these relationships, revealing how even short-term changes in symptom burden can have immediate and measurable consequences for functional capacity. The negative association between ΔPF2 and fatigue is particularly noteworthy, as fatigue is one of the most common and debilitating symptoms experienced during breast cancer treatment [42]. Fatigue reflects a complex blend of treatment-induced inflammation, metabolic alterations, sleep disturbances, and psychological strain [32]. Its strong inverse correlation with physical functioning emphasizes that fatigue is not merely a byproduct of therapy but a central determinant of whether patients can maintain their daily physical activities.

Likewise, the negative correlation between ΔPF2 and pain highlights the multifaceted role that pain plays in functional limitation. Pain can stem from tumor burden, chemotherapy-induced neuropathy, surgical sequelae, or musculoskeletal strain, and its presence can restrict mobility, reduce stamina, and amplify emotional distress [43]. Insomnia, another domain negatively associated with ΔPF2, also plays a significant role in functional deterioration. Sleep disturbances can exacerbate daytime fatigue, reduce concentration, and impair physical recovery, creating a feedback loop that magnifies functional compromise [44]. The inverse relationship between ΔPF2 and systemic side effects underscores how toxicities inherent to pharmacotherapy—such as nausea, malaise, or generalized discomfort—translate directly into diminished physical capacity. Systemic symptoms often reflect the body’s acute response to chemotherapy or targeted therapy, and their impact on physical functioning is immediate and substantial [45,46]. By visualizing these relationships in Figure 3, the analysis demonstrates the relevance of symptom monitoring as a proxy for functional decline, especially in clinical settings where objective functional assessments may not be feasible [47]. Taken together, the negative correlations depicted in Figure 3 reinforce the interconnectedness of symptom burden and functional outcomes and highlight the importance of managing symptoms proactively to preserve patient functioning during treatment. These findings also affirm the psychometric sensitivity of the EORTC instruments, which are capable of detecting multidimensional deterioration over short treatment intervals. Importantly, the visual patterns underscore that physical functioning does not decline in isolation; instead, it reflects a composite response to the cumulative symptom load experienced by patients undergoing pharmacotherapy.

## Limitations and Future Directions

This study has several limitations that should be considered when interpreting its findings. First, although conducted prospectively, the analysis was limited to QoL changes after a single cycle of pharmacotherapy, which may not capture the full trajectory of functional decline or recovery across prolonged treatment courses; future studies should incorporate longitudinal assessments across multiple cycles and into survivorship. Second, the reliance on consecutive sampling from two referral hospitals in Surabaya may limit generalizability to rural populations or healthcare settings with different resource availability, diagnostic capabilities, or cultural perceptions of symptom reporting. Third, while the EORTC QLQ-C30 and QLQ-BR23 provide comprehensive multidimensional assessments, the study did not incorporate objective functional measures, biomarkers, or clinician-reported toxicity grades, which could complement patient-reported outcomes and enable multimodal correlation analyses. Fourth, some IHC subtypes and symptom domains had incomplete or sparse data, which may have reduced statistical sensitivity for certain correlations. Future research should aim for larger, more molecularly stratified cohorts and integrate advanced analytic approaches—such as network modeling, latent class analysis, or machine-learning–based symptom clustering—to better delineate the structure and clinical implications of QoL interdependencies. Additionally, future work should evaluate how early changes in PRO correlations can predict treatment adherence, toxicity escalation, hospitalization, or long-term wellbeing, particularly in resource-limited settings where proactive symptom management may substantially improve clinical outcomes.

## Conclusion

In this prospective study of Indonesian breast cancer patients undergoing pharmacotherapy, multidimensional PROs captured through the EORTC QLQ-C30 and QLQ-BR23 provided a detailed and coherent picture of how functional and symptom domains shift during treatment. Baseline assessments revealed high functional capacity with selective symptom burdens, while correlation analyses demonstrated structured, clinically meaningful patterns in the way QoL domains changed—functional scales rising and falling together, symptom scales clustering tightly, and functional–symptom interactions consistently moving in opposing directions. These findings highlight the sensitivity and utility of validated QoL instruments in detecting early treatment-related changes and the interdependent nature of patient experiences during systemic therapy. By elucidating these correlation structures, the study underscores the importance of integrating comprehensive PROs into routine oncology practice to guide supportive care, tailor interventions, and improve patient-centered treatment strategies, particularly in resource-limited settings.

## Credit Statement

**HS:** Formal analysis, Investigation, Resources, Writing – Original Draft, Writing – Review & Editing.

**MS:** Conceptualization, Writing – Original Draft, Writing – Review & Editing, Supervision.

**EH:** Writing – Original Draft, Writing – Review & Editing, Supervision

**AA:** Writing – Original Draft, Writing – Review & Editing, Supervision

## Data Availability

All data produced in the present study are available upon reasonable request to the authors

